# Prophylaxis with tetracyclines in ARDS: Potential therapy for COVID-19-induced ARDS?

**DOI:** 10.1101/2020.07.22.20154542

**Authors:** James D. Byrne, Rameen Shakur, Joy E. Collins, Sarah Becker, Cameron C. Young, Hannah Boyce, Giovanni Traverso

## Abstract

There is an immediate need for therapies related to coronavirus disease 2019 (COVID-19), especially candidate drugs that possess anti-inflammatory and immunomodulatory effects with low toxicity profiles. We hypothesized the application of pleiotropic tetracyclines as potential therapeutic candidates. Here, we present a retrospective multi-institutional cohort study evaluating ventilatory status in patients who had taken a tetracycline antibiotic within a year prior to diagnosis of acute respiratory distress syndrome (ARDS). The primary outcomes were the need for mechanical ventilation and duration of mechanical ventilation. The secondary outcome was the duration of intensive care unit (ICU) stay. Data was evaluated using logistic regression and treatment effects regression models. Minocycline or doxycycline treatment within a year prior to ARDS diagnosis was associated with a 75% reduced likelihood for mechanical ventilation during hospital stay. Furthermore, tetracycline antibiotic therapy corresponded to significant reductions in duration of mechanical ventilation and ICU stay in ARDS patients. These data suggest tetracyclines may provide prophylactic benefit in reducing ventilatory support for ARDS patients and support further evaluation in a randomized prospective trial.

## Introduction

Severe acute respiratory syndrome coronavirus 2 (SARS-CoV-2) is the causative agent of the COVID-19 disease. One of the most significant clinical pathologies attributable to SARS-CoV-2 are its effects on the pulmonary system, with critical cases progressing to hypoxemic respiratory failure and the development of ARDS [1,2]. To date, there remains no substantive therapy for ARDS, and management continues to be largely supportive [3]. Given the immediate need for therapies related to COVID-19, especially candidate drugs which possess demonstrated anti-inflammatory and immunomodulatory effects with low toxicity profiles, we hypothesized the application of the pleiotropic tetracyclines as potential therapeutic candidates. Tetracyclines have demonstrated potent anti-inflammatory effects in addition to their antibacterial properties, including inhibition of T-cell proliferation and reduction of inflammatory cytokines, and have been used in the treatment of human immunodeficiency virus and malaria [4]. However, tetracyclines relative efficacy in ARDS patients remains undetermined. In this retrospective multi-institutional cohort study, we aimed to assess whether prophylactic use of either minocycline, doxycycline, or tetracycline could reduce the concomitant requirement for ventilatory support and duration of ICU stay among ARDS patients.

## Results

Minocycline (p = 0.037) or doxycycline (p = 0.035) treatment within a year prior to ARDS diagnosis was associated with a 75% reduced likelihood for mechanical ventilation during hospital stay (**Figure 1, A**). Similarly, treatment effects regression modeling indicated that minocycline (p = 0.004), doxycycline (p = 0.04), and tetracycline (p < 0.001) therapy corresponded to significant reductions in duration of mechanical ventilation in ARDS patients. Duration of ICU stay for patients who were previously administered minocycline (p = 0.04) or tetracycline (p = 0.014) was significantly reduced (**Figure 1, B**). Of the three tetracycline antibiotics studied, there was no significant effect in timing of administration compared to diagnosis of ARDS, except for doxycycline (**Figure 1, C**).

**Figure 1.**
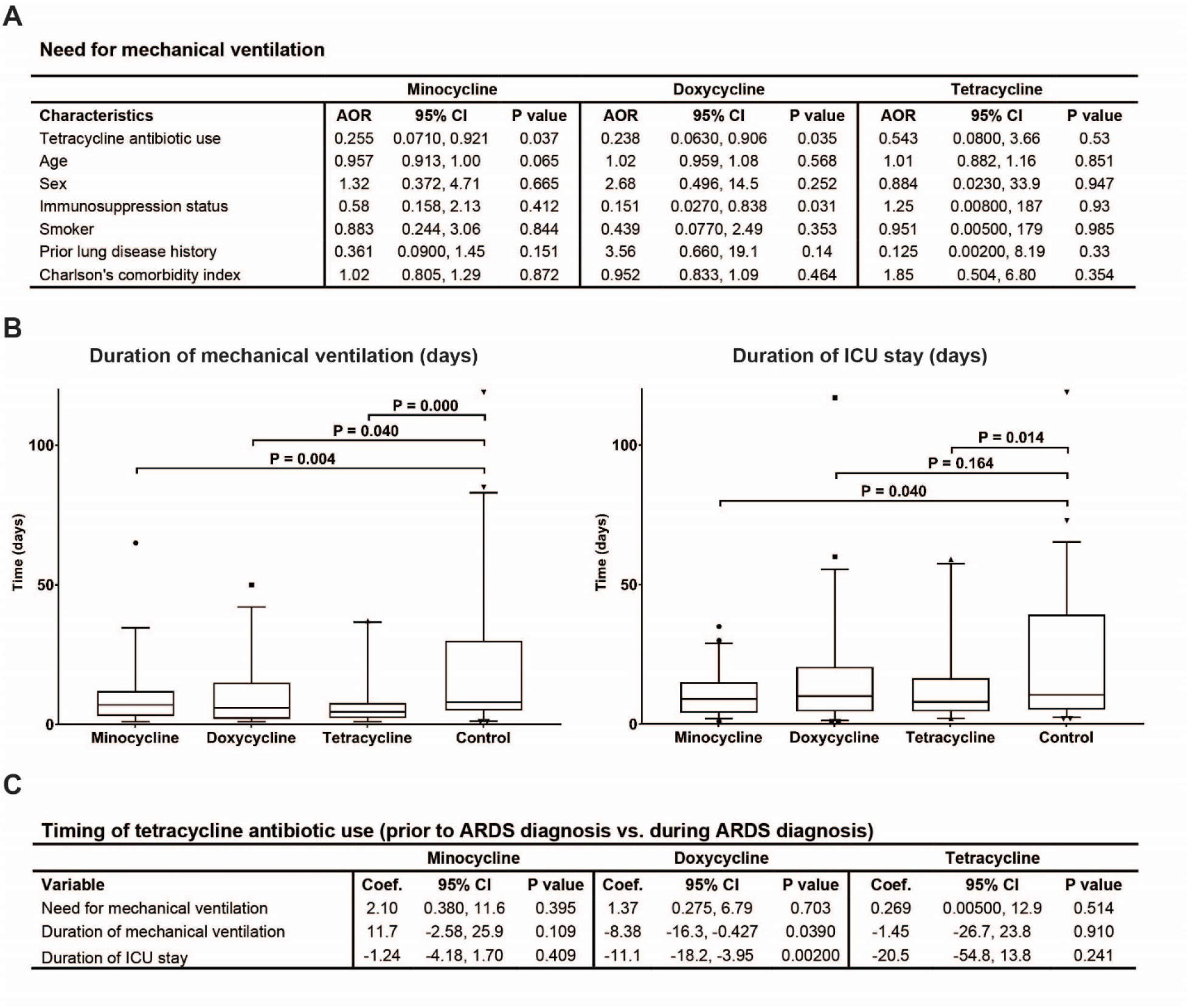
Prophylactic use of minocycline, doxycycline, or tetracycline reduces need for ventilatory support from ARDS. (A) Logistic regression of need for mechanical ventilation and treatment effects regression models. (B) Treatment effects regression models showing a reduction in the duration of mechanical ventilation and ICU stay for patients on minocycline, doxycycline, or tetracycline. (C) Treatment effects regression model for timing of the antibiotic relative to the diagnosis of ARDS. The use of minocycline, doxycycline, or tetracycline for up to a year prior to diagnosis was compared to use of the antibiotic during ARDS diagnosis, which demonstrated a treatment difference only in patients that had taken doxycycline.

## Discussion

Our results proffer the potential for tetracyclines to provide prophylactic benefit in reducing ventilatory support and duration of ICU stay for ARDS patients. Despite the retrospective nature and small sample size, we envisage implications for prophylactic tetracycline therapy in patients with ARDS secondary to COVID-19. The temporal relationship with tetracycline antibiotics’ conditioning of the immune system has been previously demonstrated in numerous clinical settings, including multiple sclerosis, rheumatoid arthritis, major depressive disorder, and inflammatory bowel disease [5-9]. Our findings are further supported by known anti-inflammatory and anti-viral effects of these antibiotics through mechanisms including downregulation of CD40 ligand on T-cells, induction of apoptosis in mast cells, and reduction in metalloproteases through zinc chelation [4]. Coronaviruses rely on metalloproteases for viral proliferation, and appear to increase mast cell proliferation in the respiratory submucosa, thus contributing to the local inflammation of lung tissue [6]. As tetracycline antibiotics are well tolerated and orally bioavailable, randomized prospective trials should be possible to further test their efficacy as a prophylactic therapy, specifically in patients at risk for the SARS-CoV-2 infection and development of ARDS.

## Methods

We performed a retrospective 20-year cohort analysis using the Partners Healthcare Research Patient Data Registry to identify patients diagnosed with ARDS to yield a total of 37,602 patients. From these patients, we then identified a subset of 49, 50, and 24 patients who had taken minocycline, doxycycline, and tetracycline, respectively, within a year prior to diagnosis of ARDS (**Supplementary Table 1**). These patients were compared to a control dataset of 49 patients diagnosed with ARDS that had not taken these antibiotics within a year of diagnosis. Analyses were performed using a logistic regression model to estimate the adjusted odds ratios (AOR) and confidence intervals (CI) to measure association between treatment (minocycline, doxycycline, or tetracycline within a year prior to ARDS diagnosis) and outcome of mechanical ventilation. Covariates including comorbid conditions, age, sex, smoking status, date of diagnosis, and immunosuppression, were adjusted in the model. Furthermore, we used treatment effects regression models to estimate the regression coefficient for the effect of minocycline, doxycycline, and tetracycline on the outcome of mechanical ventilation and duration for both mechanical ventilation and ICU stay. The effect of timing of these drugs compared to the diagnosis of ARDS were also compared. These models incorporated propensity score matching for age, sex, immunosuppression status, comorbid conditions, smoking status, and date of diagnosis. Appropriate institutional review board approval (#2020P001110) was granted by the Partners review board. Informed consent was waived due to the retrospective nature of the research. Further, all methods were performed in accordance with the relevant Partners review board guidelines and regulations.

## Data Availability

The authors declare that the data supporting the findings of this study are available within the paper and its supplementary information files.

## Acknowledgements

We would like to thank Elizabeth Limanto for her critical assessment of this work.

## Authors’ contributions

J.D.B., R.S., J.E.C., G.T. conceived and designed the study. J.D.B., R.S., J.E.C., S.B., C.C.Y., and H.B. acquired, analyzed, and interpreted the data. J.D.B., R.S., J.E.C., C.C.Y, and G.T. wrote the manuscript. G.T. supervised, reviewed the data and edited the manuscript.

## Additional Information

### Ethics approval and consent to participate

Appropriate institutional review board approval (#2020P001110) was granted by the Partners review board. Informed consent was waived due to the retrospective nature of the research.

